# Anti-spike antibody trajectories in individuals previously immunised with BNT162b2 or ChAdOx1 following a BNT162b2 booster dose

**DOI:** 10.1101/2022.02.07.22270451

**Authors:** Alexei Yavlinsky, Sarah Beale, Vincent Nguyen, Madhumita Shrotri, Thomas Byrne, Cyril Geismar, Ellen Fragaszy, Susan Hoskins, Wing Lam Erica Fong, Annalan M D Navaratnam, Isobel Braithwaite, Parth Patel, Jana Kovar, Andrew Hayward, Robert W Aldridge

## Abstract

The two most commonly-used SARS-CoV-2 vaccines in the UK, BNT162b2 (Pfizer-BioNTech) and ChAdOx1 nCoV-19 (Oxford-AstraZeneca), employ different immunogenic mechanisms. Compared to BNT162b2, two-dose immunisation with ChAdOx1 induces substantially lower peak anti-spike antibody (anti-S) levels and is associated with a higher risk of breakthrough infections. To provide preliminary indication of how a third booster BNT162b2 dose impacts anti-S levels, we performed a cross-sectional analysis using capillary blood samples from vaccinated adults (aged ≥18 years) participating in Virus Watch, a prospective community cohort study in England and Wales. Blood samples were analysed using Roche Elecsys Anti-SARS-CoV-2 S immunoassay. We analysed anti-S levels by week since the third dose for vaccines administered on or after September 1, 2021 and stratified the results by second dose vaccine type (ChAdOx1 or BNT162b2), age, sex and clinical vulnerability. Anti-S levels peaked at two weeks post-booster for BNT162b2 (22,185 U/mL; 95%CI: 21,406-22,990) and ChAdOx1 second dose recipients (19,203 U/mL; 95%CI: 18,094-20,377). These were higher than the corresponding peak antibody levels post-second dose for BNT162b2 (12,386 U/mL; 95%CI: 9,801-15,653, week 2) and ChAdOx1 (1,192 U/mL; 95%CI: 818-1735, week 3). No differences emerged by second dose vaccine type, age, sex or clinical vulnerability. Anti-S levels declined post-booster for BNT162b2 (half-life=44 days) and ChAdOx1 second dose recipients (half-life=40 days). These rates of decline were steeper than those post-second dose for BNT162b2 (half-life=54 days) and ChAdOx1 (half-life=80 days). Our findings suggest that peak anti-S levels are higher post-booster than post-second dose, but that levels are projected to be similar after six months for BNT162b2 recipients. Higher peak anti-S levels post-booster may partially explain the increased effectiveness of booster vaccination compared to two-dose vaccination against symptomatic infection with the Omicron variant. Faster waning trajectories post third-dose may have implications for the timing of future booster campaigns or four-dose vaccination regimens for the clinically vulnerable.

## Introduction

Breakthrough SARS-CoV-2 infections in individuals who have received two doses of a COVID-19 vaccine are now routinely reported [1,2]. These are partially attributed to anti-spike antibody (anti-S) waning within several months of receiving the second dose and to the emergence of new variants [3–6]. Consequently, the UK government initiated a booster vaccination campaign, initially targeting people over the age 50, adults deemed clinically vulnerable to severe COVID-19 outcomes, care home residents, frontline health workers, and household contacts of immunocompromised individuals [7]. This was subsequently expanded to the entire adult population following the emergence of the immune-evasive Omicron (B1.1.529) variant [8].

The two most commonly-used vaccines in the UK, BNT162b2 (Pfizer–BioNTech) and ChAdOx1 nCoV-19 (Oxford–AstraZeneca), employ different immunogenic mechanisms. Compared to BNT162b2, two-dose immunisation with ChAdOx1 induces substantially lower peak antibody levels and is associated with a higher risk of breakthrough infections [9].

To provide preliminary indication of how a third booster BNT162b2 dose impacts anti-S levels, we performed a cross-sectional analysis using capillary blood samples from vaccinated adults (aged ≥18 years) participating in Virus Watch, a prospective community cohort study in England and Wales [10]. Virus Watch received ethical approval from the Hampstead NHS Health Research Authority Ethics Committee (20/HRA/2320).

## Methods

Sera were tested for antibodies to the S1 subunit of the wild-type SARS-CoV-2 spike protein (range 0.4–25 000 units per mL [U/mL]) using the Elecsys Anti-SARS-CoV-2 S immunoassay, and for antibodies to the full-length nucleocapsid protein as a proxy for prior SARS-CoV-2 infection (specificity 99.8% [99.3–100]) using the N electro-chemiluminescent immunoassay (Roche Diagnostics, Basel, Switzerland). Serological results were linked with demographic and clinical information collected at registration and with weekly self-reported vaccination status.

We analysed anti-S levels by week since the third dose for vaccines administered on or after September 1, 2021 and stratified the results by second dose vaccine type (ChAdOx1 or BNT162b2), age, sex and clinical vulnerability (defined in Tables S2 and S3). Individuals who had previously submitted blood samples that were seropositive for anti-Nucleocapsid antibodies were excluded. We also compared anti-S levels post-second dose to those post-booster. To avoid ceiling effects due to samples falling above the 25,000 U/mL threshold, for each week we computed the median value of log anti-S levels as an approximation of the geometric mean. This median value remains accurate as on any given week over 50% of the post-booster samples had antibody levels below the 25,000 U/ml threshold. Median absolute difference in the log space was then used to approximate the standard error for calculation of 95% confidence intervals around the medians [11]. Anti-S level half-lives were obtained by performing linear regression on the log values of the weekly median estimates.

## Results

In the post-booster cohort, 4,682 adults submitted one or more valid samples between August 18, 2021 and January 6, 2022 (7,406 total samples). In the post-second dose cohort, 8,680 adults submitted one or more valid samples between July 1, 2021 and January 6, 2022 (24,271 total samples). Table 1 in the supplementary material appendix shows demographic and clinical characteristics of the two cohorts.

Anti-S levels peaked at two weeks post-booster for BNT162b2 (22,185 U/mL; 95%CI: 21,406-22,990) and ChAdOx1 second dose recipients (19,203 U/mL; 95%CI: 18,094-20,377). These were higher than the corresponding peak antibody levels post-second dose for BNT162b2 (12,386 U/mL; 95%CI: 9,801-15,653, week 2) and ChAdOx1 (1,192 U/mL; 95%CI: 818-1735, week 3) (Figure 1). As 41.3% of BNT162b2 and 33.6% of ChAdOx1 peak-level samples equalled or exceeded the 25,000 U/mL measurement threshold (Supplementary Figure S1), the width of the corresponding confidence intervals may be underestimated and the lack of overlap should be interpreted cautiously; median estimates are unaffected. No differences emerged by second dose vaccine type, age, sex or clinical vulnerability (Figure 1 and Figure S2).

**Figure 1.**
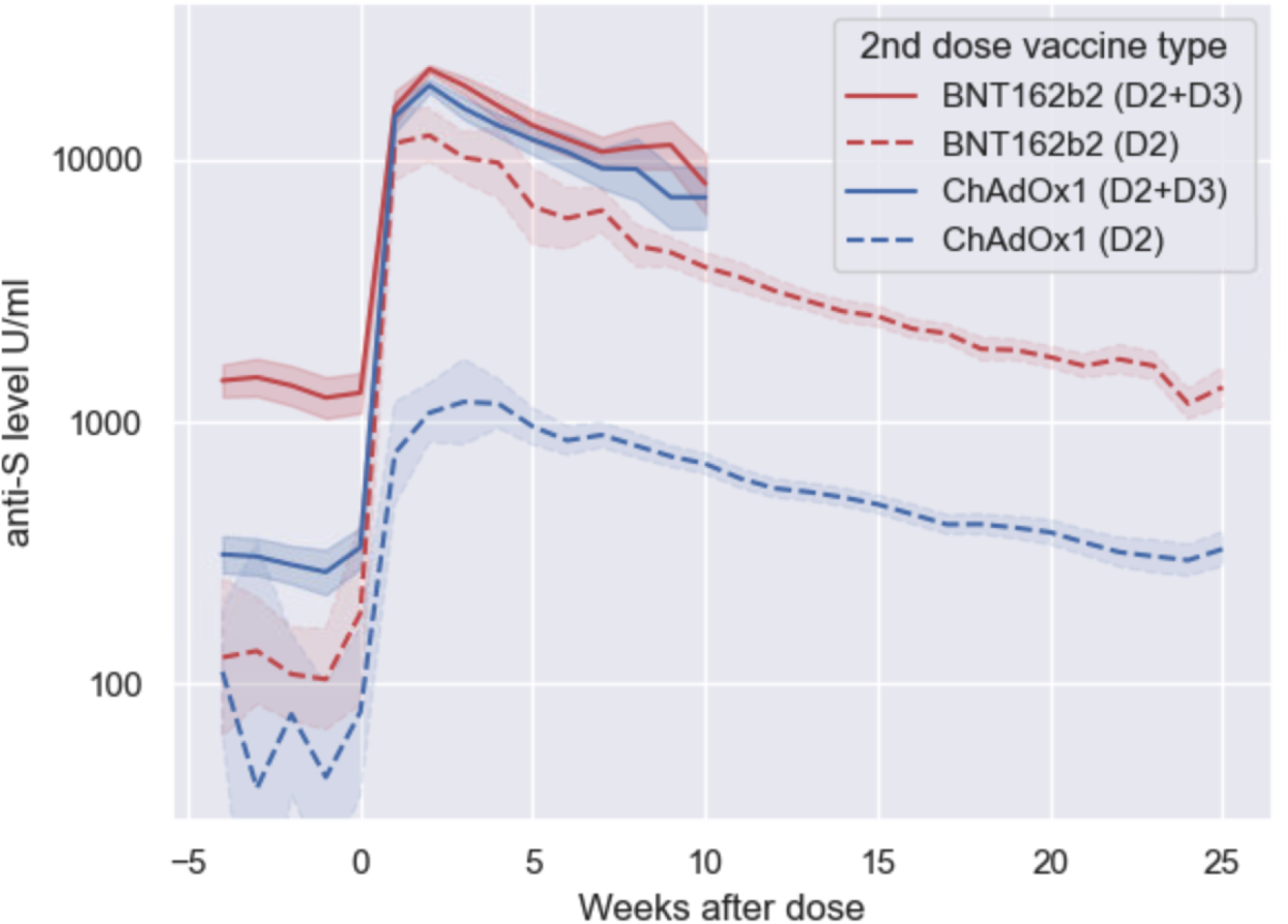
Anti-S levels (U/mL) over time since BNT162b2 booster dose (D2+D3) and second vaccine dose (D2) amongst N-seronegative individuals by second dose vaccine type.

Anti-S levels declined post-booster for BNT162b2 (half-life=44 days) and ChAdOx1 second dose recipients (half-life=40 days). These rates of decline were steeper than those post-second dose for BNT162b2 (half-life=54 days) and ChAdOx1 (half-life=80 days). For BNT162b2 second dose recipients, median anti-S levels at 26 weeks were predicted to be 1,487 U/m post-booster compared to 1,285 U/mL at 26 weeks after the second dose.

## Discussion

Our findings suggest that peak anti-S levels are higher post-booster than post-second dose, but that levels are projected to be similar after six months for BNT162b2 recipients. No differences in post-booster antibody levels emerged by second dose vaccine type. This finding contrasts with antibody responses post-second dose, which were substantially lower for ChAdOx1 than BNT162b2 recipients. The magnitude and trajectory of post-booster anti-S response was similar across age groups and by clinical vulnerability status. Higher peak anti-S levels post-booster may partially explain the increased effectiveness of booster vaccination compared to two-dose vaccination against symptomatic infection with the Omicron variant.

We measured antibody levels using an assay developed against earlier strains of COVID-19 and did not measure neutralising antibody levels. Due to immune escape, higher circulating antibody levels are likely to be needed to protect against Omicron variant infection compared to previous strains. It should also be noted that immune mechanisms other than circulating antibody levels such as T and B cell memory responses are likely to partially mediate protection against severe disease [12,13].

Anti-S levels also appeared to wane faster following the booster dose; however, ten weeks after vaccination these remained above previously-estimated thresholds for breakthrough infection with the Delta variant due higher peak levels [9]. Thresholds for breakthrough infection with the Omicron variant are currently unknown. Faster waning trajectories post third-dose may have implications for the timing of future booster campaigns or four-dose vaccination regimens for the clinically vulnerable. Despite a faster waning trajectory, booster vaccination appears to substantially enhance anti-S levels - and likely consequent protection against symptomatic infection and severe outcomes - to a uniform degree across age and clinical risk groups.

ACH serves on the UK New and Emerging Respiratory Virus Threats Advisory Group. All other authors declare no competing interests. The Virus Watch study is supported by the MRC Grant Ref: MC_PC 19070 awarded to UCL on 30 March 2020 and MRC Grant Ref: MR/V028375/1 awarded on 17 August 2020. The study also received $15,000 of Facebook advertising credit to support a pilot social media recruitment campaign on 18th August 2020. This study was also supported by the Wellcome Trust through a Wellcome Clinical Research Career Development Fellowship to RA [206602]. SB and TB are supported by an MRC doctoral studentship (MR/N013867/1). The funders had no role in study design, data collection, analysis and interpretation, in the writing of this report, or in the decision to submit the paper for publication

## Data Availability

We aim to share aggregate data from this project on our website and via a “Findings so far” section on our website - https://ucl-virus-watch.net/. We will also be sharing individual record level data on a research data sharing service such as the Office of National Statistics Secure Research Service. In sharing the data we will work within the principles set out in the UKRI Guidance on best practice in the management of research data. Access to use of the data whilst research is being conducted will be managed by the Chief Investigators (ACH and RWA) in accordance with the principles set out in the UKRI guidance on best practice in the management of research data. We will put analysis code on publicly available repositories to enable their reuse.

