## Supplementary material for "Anti-spike antibody trajectories in individuals previously immunised with BNT162b2 or ChAdOx1 following a BNT162b2 booster dose"

**Table S1.** Demographic and clinical characteristics of the post-second dose and post-booster cohorts.

| | Third (booster) dose cohort | Second dose cohort |
|---|---|---|
| **Number of participants** | 4,682 | 8,680 |
| **Age group** | | |
| 18-64 | 1,971 (42%) | 4,500 (52%) |
| 65+ | 2,711 (58%) | 4,180 (48%) |
| **Sex** | | |
| Female | 2,719 (58%) | 5,004 (58%) |
| Male | 1,957 (42%) | 3,666 (42%) |
| Other/Missing | 6 (0.1%) | 10 (0.1%) |
| **Clinical vulnerability** | | |
| Clinically extremely vulnerable | 715 (15%) | 1,207 (14%) |
| Clinically vulnerable | 1,293 (28%) | 2,352 (27%) |
| Not clinically vulnerable | 2,674 (57%) | 5,121 (59%) |
| **2nd dose vaccine type** | | |
| BNT162b2 | 1,871 (40%) | 3,119 (36%) |
| ChAdOx1 | 2,778 (59%) | 5,449 (63%) |
| Other/Missing | 33 (0.7%) | 112 (1.3%) |

**Figure S1.** Proportion of post-booster samples exceeding 25,000 U/mL for second dose BNT162b2 and ChAdOx1 recipients.

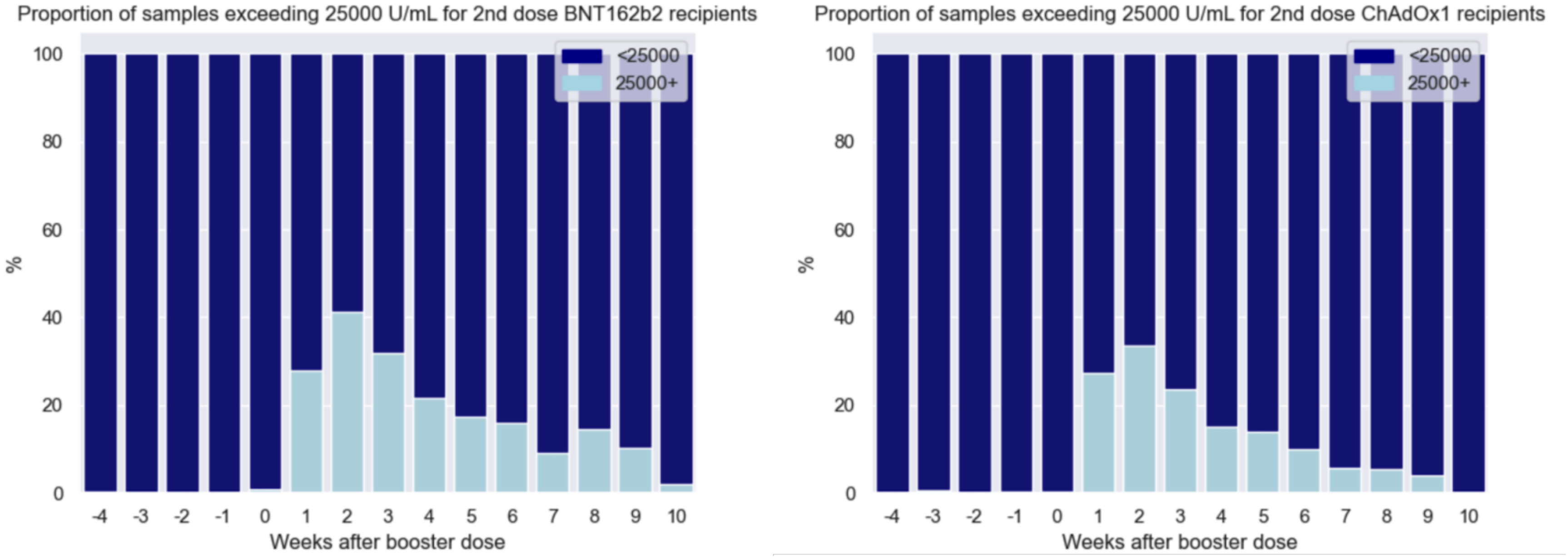

**Figure S2.** Anti-S levels (U/mL) over time since BNT162b2 booster dose amongst N-seronegative individuals by second dose vaccine type and age, sex and clinical risk group.

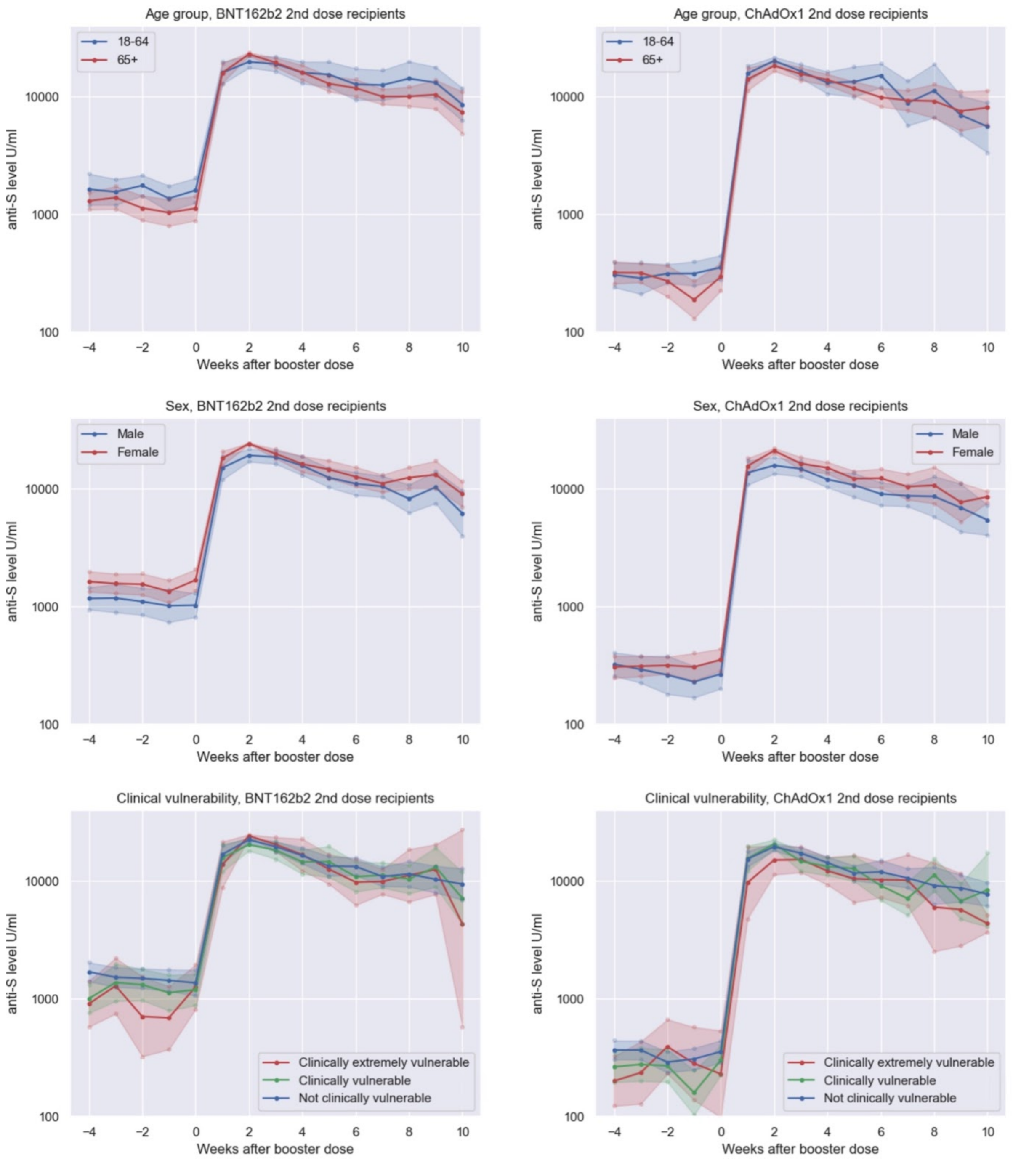

**Table S2.** Clinically extremely vulnerable classification.

Individuals were categorised as extremely clinically vulnerable using criteria set out by Public Health England and the Department of Health and Social Care as part of the guidance for shielding (https://www.gov.uk/government/publications/guidance-on-shielding-and-protecting-extremely-vulnerable-persons-from-covid-19), which were adapted in line with clinical variables collected through the Virus Watch baseline survey, as follows:

| Clinically extremely vulnerable (CEV) criteria as per PHE/DHSC | Inclusion in Virus Watch CEV definition |
|---|---|
| Solid organ transplant recipients | Included |
| Cancer undergoing active chemotherapy | Included |
| Cancers undergoing radical radiotherapy | All radiotherapy included (radical radiotherapy was not ascertained) |
| Cancer of blood or bone marrow | Included |
| Immunotherapy or antibody treatments for cancer | Included |
| Targeted cancer therapies affecting the immune system | Included |
| Bone marrow or stem cell transplant in last 6 months or still taking immunosuppressive drugs | Included |
| Severe respiratory conditions including all cystic fibrosis, severe asthma and severe chronic obstructive pulmonary disease (COPD) | Included |
| Rare diseases that significantly increase the risk of infections (such as severe combined immunodeficiency (SCID), homozygous sickle cell disease) | Included |

| | |
|---|---|
| Immunosuppressive therapies sufficient to significantly increase risk of infection | Included |
| Problems with spleen, including splenectomy | Included |
| Down's syndrome | Not included in CEV as not distinguished from other learning disabilities. |
| Chronic kidney disease Stage 5 or on renal dialysis | All CKD was included (stage was not ascertained) |
| Pregnancy with significant heart disease | Included |
| Others classified as clinically extremely vulnerable | Included |

**Table S3.** Clinically vulnerable classification.

Individuals were categorised as clinically vulnerable (CV) using criteria set out by the Joint Committee on Vaccination and Immunisation (https://www.gov.uk/government/publications/priority-groups-for-coronavirus-covid-19-vaccination-advice-from-the-jcvi-30-december-2020), excluding those who met the superseding clinically extremely vulnerable (CEV) criteria. Clinical vulnerability criteria were adapted in line with clinical variables collected through the Virus Watch baseline survey, as follows:

| **Clinically vulnerable (CV) criteria as per JCVI** | **Inclusion in Virus Watch CV definition** |
|---|---|
| chronic respiratory disease, including chronic obstructive pulmonary disease (COPD), cystic fibrosis and severe asthma | Included, except those that met CEV criteria |
| chronic heart disease (and vascular disease) | Included |

| | |
|---|---|
| chronic kidney disease | Included, except those that met CEV criteria |
| chronic liver disease | Included |
| chronic neurological disease including epilepsy | Included |
| Down’s syndrome | Included as part of broader learning disabilities |
| Severe and profound learning disability | All learning disabilities included (severity was not ascertained) |
| Diabetes | Included |
| Solid organ, bone marrow and stem cell transplant recipients | Not included (included in CEV) |
| People with specific cancers | Included, except those that met CEV criteria |
| Immunosuppression due to disease or treatment | Included, except those that met CEV criteria |
| Asplenia and splenic dysfunction | Not included (included in CEV) |
| Morbid obesity | Included |
| Severe mental illness | Included |
